## Appendix for "Using genetic data to identify transmission risk factors: statistical assessment and application to tuberculosis transmission"

### A Appendix

#### A.1 Methods

##### A.1.1 Simulation Settings

| Setting | True OR | Clusters per Sim. | Samp. Prop. | Samp. Window |
|---|---|---|---|---|
| 0.3 | 0.25 | 50 | 0.16 | 3 |
| 0.57 | 0.52 | 50 | 0.16 | 3 |
| 1 | 1 | 50 | 0.16 | 3 |
| 1.75 | 1.94 | 50 | 0.16 | 3 |
| 3 | 3.53 | 50 | 0.16 | 3 |
| Increase Sample Size | 1.94 | 100 | 0.16 | 3 |
| Double Sampling Density | 1.94 | 25 | 0.32 | 3 |
| Quad Sampling Density | 1.94 | 13 | 0.64 | 3 |
| Increase Sampling Window | 1.94 | 35 | 0.16 | 7 |

**Table A1.** Simulation settings used in this study.

##### A.1.2 `TransPhylo` parameters and priors

| Parameter | Defn. | Prior | Value/Prior Median & 95% Quantiles |
|---|---|---|---|
| $\pi$ | Sampling prop. | Beta(1,19) | 0.036 (0.0014, 0.18) |
| $r$ | Reproduction number | Exp(1) | 0.68 (0.023, 3.70) |
| $p$ | No effect | NA | 0.5 |
| $N_{eg}$ | W/in-host ESS | Exp(1) | 0.68 (0.023, 3.70) |
| $\alpha_{gen}$ | Gen. time shape | NA | 10 |
| $\beta_{gen}$ | Gen. time scale | NA | 0.1 |
| $\alpha_{samp}$ | Samp. time shape | NA | 10 |
| $\beta_{samp}$ | Samp. time scale | NA | 0.1 |

**Table A2.** `TransPhylo` parameters and priors

Note that we chose the sampling distributions to be the same as the generation time distributions. We felt this was plausible as it reflects our assumption that individuals in their latent stage would not be detected in our sampling regime. We chose to have the mean generation time be 1 year, based on our choices for the mean latent period (9 months) and the mean infectious period (3 months). Treating the mean generation time as the sum of the mean latent and mean infectious periods is a reasonable choice, see previous work on this topic by Svennson and Champredon et al. for more details [33, 34]. Moreover, our mean generation time is consistent with published estimates for the serial interval for tuberculosis of 0.57 to 1.65 years. [35] See below for rationales behind our choices for the mean latent and infectious periods. The sampling prior was chosen based on early empirical results from simulations where we sampled only from the last three years in the sampling regime described in the main methods, which is meant to resemble the sampling regime in our study setting. The same parameters were used for both simulated and real data.

##### A.1.3 nosoi Parameters

For each simulated individual, the latent period duration, during which the individual could neither leave nor transmit infection to others, was drawn from a gamma distribution with shape parameter 6.75, rate parameter .75 so that the mean was 9 months. Incubation periods for tuberculosis are estimated to be between a few months to two years in duration, with limited instances of longer duration [36]. Based on this, the latent period for each simulated individual, during which the individual could neither leave nor transmit infection to others, was drawn from a gamma distribution with shape parameter 6.75, rate parameter .75 so that the majority of individuals had latent period of less than 2 years, with a mean duration of 9 months. The latent period was considered to be over at the first time step greater than or equal to the latent period length. For simplicity, individuals who are infected, but never progress to infectious TB are ignored. The probability of exiting the simulation was drawn from a beta distribution with $\alpha = 1/0.11$ and $\beta = 3/0.11$ so that the mean probability was 0.25 (after the latent period was over). A mean probability of 0.25 translates into an average of three months of time during which an individual is actively infectious. For this distribution, we assumed that individuals become infectious when they start coughing and become non-infectious shortly after diagnosis and treatment. We previously found that 50% of TB diagnosis and treatment occurred within 1 month and 80% within 2 months of cough onset among people with TB in Botswana. [37] We assumed a long tail for delayed diagnosis and treatment for TB. [38] We used a discrete population structure with two locations (A and B) to create our two classes of cases. Individuals had the chance to move exactly once from their starting location, and the probability of moving from A to B was 0.53, while the probability of moving from B to A was 0.47. This resulted in (in general) 53% of cases staying in location B, while 47% of cases stayed in location A. These numbers were chosen to reflect the empirical proportion of those living with HIV found in our data. Number of contacts per time step was drawn from a normal distribution with mean 20 and standard deviation 5. A contact survey found people in Zambia and South Africa found participants had about four close contacts per day [39]. A meta-analysis of contact surveys found average daily contacts globally was about 9 contacts per day [40]. We would expect that in a month, individuals see the same people multiple times, so that daily contacts don't translate to monthly contacts. This is hard to quantify rigorously, but we felt an average of 20 unique contacts a month was not unrealistic. In practice, the number of contacts mostly controls the size of the outbreak, because new infections are not dependent on current infections in our simulations. The normal random variable was then rounded and the absolute value used as the number of contacts. The basic reproduction number was chosen to be 1.18, that is, on average, individuals infect 1.18 other individuals. This number has been shown to vary substantially in different settings, ranging from 0.24 to 4.3 in previous studies. [35] We chose the reproduction number of 1.18 to mimic a disease which is spreading steadily but where the slope of incidence is not growing incredibly quickly, which reflects the situation of TB in high prevalence countries such as Botswana. We can write the basic reproduction number as a function of the average total number of contacts during the infectious period multiplied by the probability of transmission given contact. In mathematical terms:

$$\begin{aligned}\texttt{Basic Reproduction Number} = {} & \text{Average Total Contacts}* \\ & (0.47 * P(\texttt{Transmission|Contact in A})+ \\ & (0.53 * P(\texttt{Transmission| Contact in B})).\end{aligned}$$

Thus, probability of transmission given contact for each location was calculated beginning from a starting point of having the base reproduction number be 1.18, the mean number of contacts be 20, and the mean number of actively infectious time steps be 3. The exact values changed between simulation settings, depending on the desired ratio for probability of transmission given contact for the two classes of individuals. We chose this reproduction number to mimic a disease which is spreading steadily but where the slope of incidence is not growing incredibly quickly, which we think reflects the situation of TB in high prevalence countries such as Botswana.

##### A.1.4 BCa Bootstrap

We attempted using BCa Bootstrap as an additional step in the TP + GLM pipeline for the primary simulation setting where the probability of transmission given contact was 3.02 times higher for people living without HIV compared to people with HIV [41]. We found it did not produce note-able improvements in coverage, and chose not to pursue studying it further with other simulation settings.

#### A.2 Results

##### A.2.1 Monte Carlo estimates of the true odds ratios

| Setting | Point Estimate | Standard Error | 99% CI | Value Used |
|---:|---:|---:|---:|---:|
| 0.3 | 0.2487 | 0.00066 | [0.2473, 0.2499] | 0.25 |
| 0.57 | 0.5205 | 0.0013 | [0.5178, 0.5232] | 0.52 |
| 1 | 1 | NA | NA | 1 |
| 1.75 | 1.9381 | 0.00176 | [1.9345, 1.9415] | 1.94 |
| 3 | 3.5298 | 0.0014 | [3.5267, 3.5327] | 3.53 |

**Table A3.** Monte Carlo estimates for the true Odds Ratios of probability of being an infection source used in simulations. Value Used denotes the value used in the simulations when calculating frequentist metrics.

Note that in cases where there were two possible values for the 100ths decimal place of the true odds ratio of interest, we found there was little meaningful difference between using one value versus the other in terms of overall results (analysis not shown). We did not use Monte Carlo estimates for the "1" setting, as the true odds ratio is one in this case.

##### A.2.2 Sensitivity and Specificity of TransPhylo Infection Source Labels

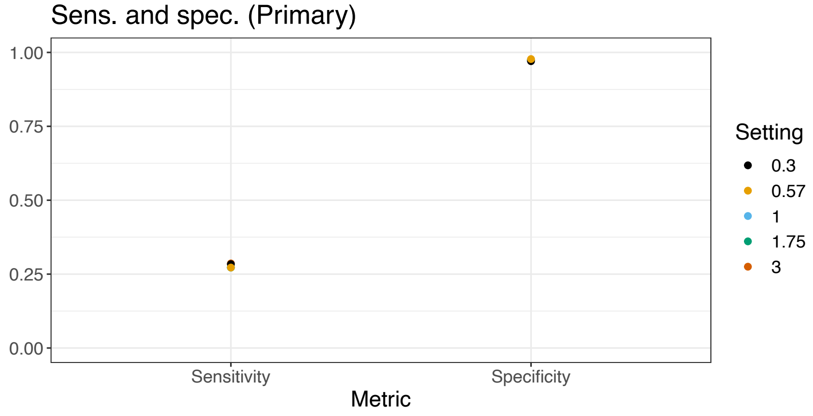


**Fig A1.** Point estimates of sensitivity and specificity of identifying infection sources based on `TransPhylo` estimates. Settings refer to simulation settings from the primary simulations.

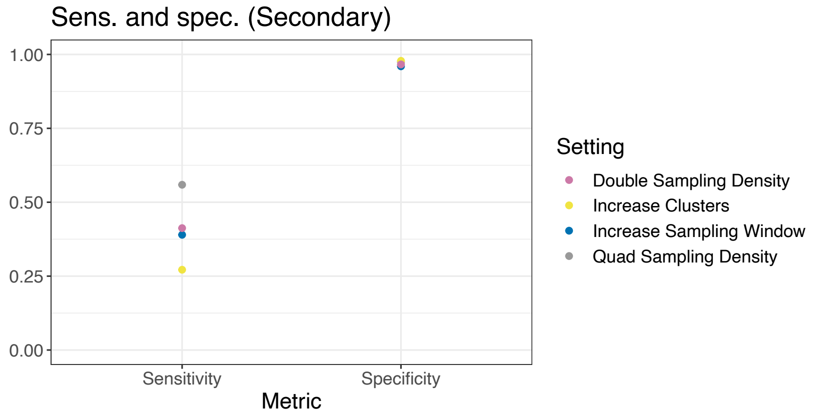


**Fig A2.** Point estimates of sensitivity and specificity of identifying infection sources based on `TransPhylo` estimates. Settings refer to simulation settings from the secondary simulations.

While we cannot assess sensitivity and specificity of `TransPhylo` on real data, we can at least look at the distribution of probabilities of being an infection source to see how robust our results are to changes in the threshold probability used to assign labels. The distribution of probabilities of being an infection source was likewise bi-modal in the real data analysis. In the 5 SNP data set, a threshold of 0.4 instead of 0.6 resulted in a single additional infection source, a threshold of 0.8 resulted in 5 fewer infection sources. In the 10 SNP data set, a threshold of 0.4 results in 17 (4% of study participants) new infection sources, a threshold of 0.8 results in 9 (2.4% of study participants) fewer infection sources.

#### A.3 Sensitivity analysis for TransPhylo labels

The table below shows how frequentist metrics for the TP + GLM pipeline change in the "1.75" setting when hosts are assigned to be infection sources when their probabilities of being an infection source are 0.4, 0.5, 0.6 (reference used in main paper) and 0.7.

| Coverage | Prop. Reject | Percent Bias | MCIW | Method | Cutoff |
|---|---|---|---|---|---|
| 0.78 | 0.54 | -0.19 | 1.29 | TP + GLM | 0.4 |
| 0.82 | 0.54 | -0.18 | 1.34 | TP + GLM | 0.5 |
| 0.85 | 0.56 | -0.15 | 1.42 | TP + GLM | 0.7 |
| 0.86 | 0.54 | -0.16 | 1.38 | TP + GLM | 0.6 (Default) |

**Table A4.** Frequentist metrics of the TP + GLM pipeline in the 1.75 primary simulation setting when hosts are labeled as an infection source based on varying cutoffs for the probability of being an infection source. For 0.4, any host with a probability of being an infection source of 0.4 or higher was labeled an infection source. In the main results, the cutoff used was 0.6. Here we alter this cutoff to 0.4, 0.5, or 0.7.

The figure below shows how sensitivity and specificity for `TransPhylo` generated labels changed under these different cutoff values.

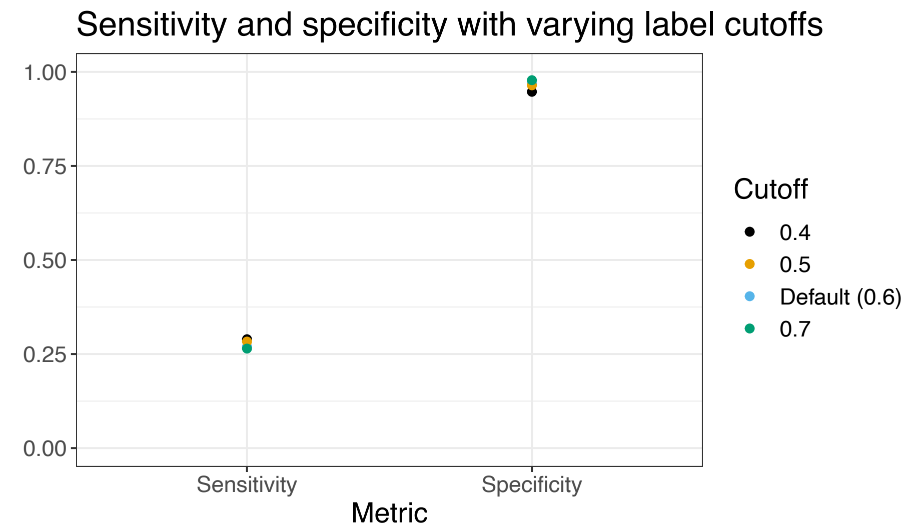


**Fig A3.** Point estimates of sensitivity and specificity of identifying infection sources based on `TransPhylo` estimates. Cutoffs refer to the varying cutoffs for the probability of being an infection source. For 0.4, any host with a probability of being an infection source of 0.4 or higher was labeled an infection source. In the main results, the cutoff used was 0.6. Here we alter this cutoff to 0.4, 0.5, or 0.7.
